# Searching the Sigmoid-type trend in Lock Down period covid19 data of India and its different states

**DOI:** 10.1101/2020.04.25.20079624

**Authors:** Supriya Mondal, Sabyasachi Ghosh

**Affiliations:** Aadarsh Nursing Institute, Raipur, Chhattisgarh 492015, India; Indian Institute of Technology Bhilai, GEC Campus, Sejbahar, Raipur 492015, Chhattisgarh, India

## Abstract

We have worked on imported covid19 cases of India, detected after declaration of lock down. They are marked as hidden travelers, who are detected later during 1-2 weeks period of lock down. We have investigated the impact of those travelers on selective 15 states, having large number cases. Among them 5 states have faced a noticeable effect on their data profile, although its overall effect in country level is quite negligible. Searching the Sigmoid trends of all 15 states, we have identified 3 categories depending upon latest data of new case with respect to the peak value of Sigmoid functions. Based on our optimistic predicted curves, ongoing lock down period might be enough for getting marginal values of new cases for KL, HR, JK, TN and AP but the reamining of 15 states might need a third phase of lock down or alternative preventive measures.

## 1 Introduction

Entire world is almost stopped by ongoing covid19 infection, which is declared as pandemic disease by World Health Organization (WHO) [1]. Understanding its epidemiological status in individual country and states might be considered as an important topic in present situation. This article is focused on India and its some selective states, having large number of covid19 cases. This kind of epidemiological study for India is started by few research groups. [2, 3, 4, 5], which should be rapidly grown by other theoretical groups for better and critical understanding. In that context, our earlier work [3, 4] have attempted to identify the time zone, from where an exponential growth almost started in India as well as its different states. Considering the last slope parameter in logarithm scale, transformation of exponential to Sigmoid function [7] is sketched for the different states and they are compared with respect to existing number of Beds in Govt. hospitals. After those initial investigations, present work is attempted to zoom in the imported covid19 data of India [6] and its connection with Sigmoid nature of updated data (till 18th April). This attempt has tried to understand imported to local transmission through different possible redistribution of existing data.

The article is organized as follows. In next section (2), we initiated the discussion on imported data through different graphical representations. Here we have observed that imported covid19 data does not immediately vanished after lock-down, rather it takes some time to be disappeared. Here, we have noticed how this imported data is gradually transformed to zero during lock-down period. Then in Sec. (3), we have identified 5 states, which face a noticeable impact from this imported data. While in Sec. (4), 10 more states, having ignorable impact of imported data, are discussed. In both sections (3) and (4), the Sigmoid trend of data are analyzed and based on that pattern, we classified the 15 states into 3 gross categories. At the end, we have summarized the study with extracting interpretation in Sec. (5).

### 2. Probing from imported to Local transmission data

Except China, for all other countries in the globe, the Covid19 is an imported disease. Then imported person spread this infection to others until diagnosed as positive. This can be visualized through the variation of ratio between imported and (imported + non-imported) or all cases of any country by increasing the days. The pattern should be as follows. In the beginning, the ratio remain one and later it becomes less than one. It reflects that first slot of all cases are imported candidate and later the local transmitted candidates came into the picture, for which the ratio transformed from one to less than one.

From covid19 data of India [6], we have identified the new cases and total cases, who came from abroad or other states to their own state. we can collectively call them as imported data. For understanding our terminology of new and total case, we mean that new case is the no. of covid19 cases detected in each day, whereas the total cases is basically cumulative of new cases against time (day) axis. The new case and total cases imported data are sketched by green triangles and blue squares in left panel of Fig. (1). From day 1 (30^th^ Jan, 2020) to day 55 (25^th^ March, 2020), all imported cases have travel history from abroad (except 2-5 candidates) but from 25^th^ March to 8^th^ April, we get a large number of imported cases, who did not come from abroad, rather came from other states. The brown circles in the left panel of Fig. (1) denotes the imported from abroad cases, which is not equal to total imported cases.

**Figure 1:**
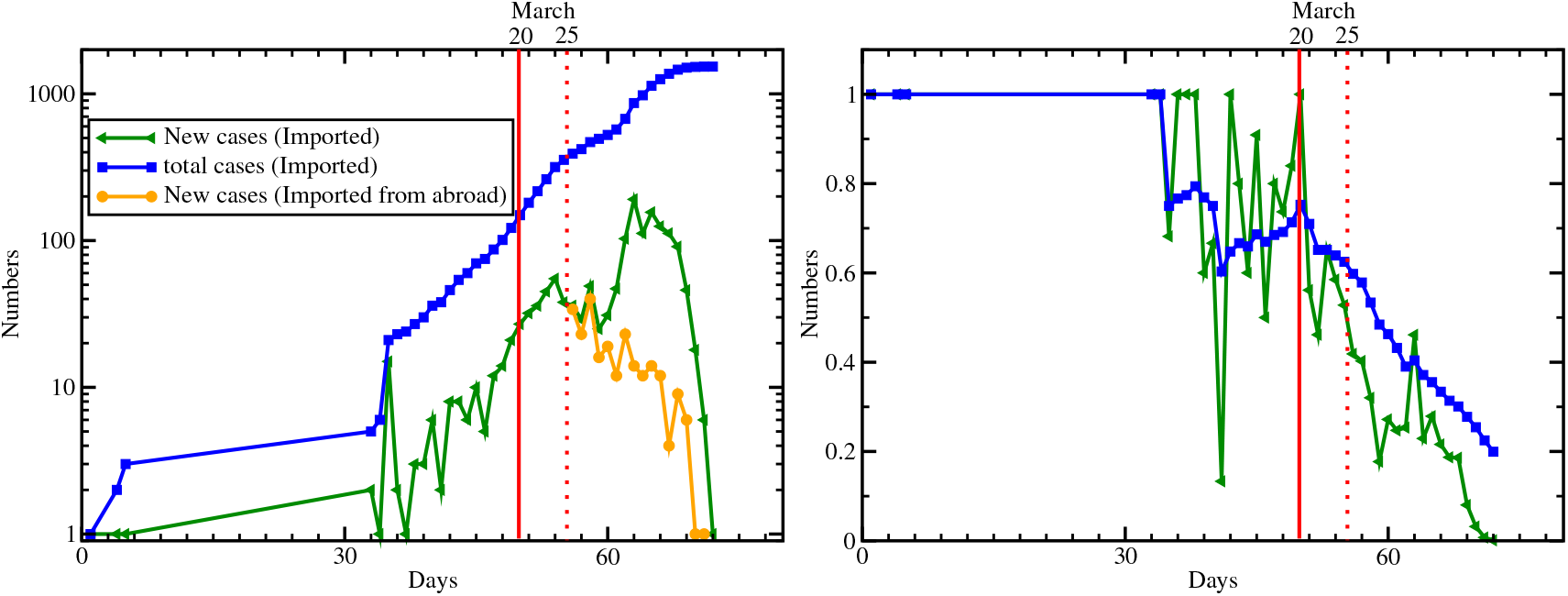
Left panel : Imported new cases (green triangles), total cases (blue squares) and imported from abroad (new) cases only (brown circles). Right panel : ratio between imported and all for new cases (green triangles) and total cases (blue squares). Red solid and dotted line denote 20^th^ and 25^th^ March (2020) respectively.

Next in the right panel of Fig. (1), we have drawn the ratio of imported to all by green triangles and blue squares for new and total cases. Till 3-4 March, total case remain lower than 10 and all are imported cases. Therefore, we see ratio 1 for a long period in the right panel of Fig. (1). After that, the ratio started to reduce, which implies that imported fellow now have started to infect others (may be his/her family members or close contact candidates). The new case ratio is too fluctuating, whereas total case ratio is little stable within 0.6-0.8. Interestingly, ratio first decreases to 0.6 and then increases to 0.75 at 20^th^ March, when airlines and rail transport are declared for shutting down in India. After that the ratio constantly decreases as expected due to continuation of lock down. Vertical red solid line indicates the date of 20^th^ March and red dotted line denotes the 25^th^ March, from when lock-down in whole country started.

The motivation of present investigation comes from an interesting observations - after announcing the lock down from 25th March, we still can find covid-19 positive cases who had travel history [6]. It means that they traveled before 25th March but they are detected as a positive case later on. Since India Govt. shut down airlines and railway services from 20th March, 2020, so most probably their journey date may be before 20th March.

In Fig. (2), we have plotted the number of covid +ve cases, who came from abroad (black squares) and from outside state to their own state (red circles). The data in the curves is for entire country and plotted against day-axis, which started from day 56 (25th March, 2020) and roughly extended to 2 weeks. Corresponding total data for different states and India are documented in Table (1). Here, initial point *t* = 0 corresponds to 30^th^ January therefore the data from 25^th^ March has been started from day 56. Now we see that total 240 traveler from abroad and 938 traveler from other states are detected as +ve cases after 25th March, although their travel journey probably happened before 25th March or most probably before 20th March. It means that their covid19 symptom take time to reveal. Since this no. is vanished around 8th April, shown by Fig. (2), so we may accept the fact that the 2-3 weeks after lock down/shutting down the transportation might be considered as affected period, during when the hidden covid19 carrier may reveal their symptoms. Let us called them as HTs (hidden travelers) for our convenience.

**Table 1:** Numbers of covid +ve cases, who came from abroad (second row) and from outside state (third row), marked as *Imported* (*from abroad*) and *State to state traveler* respectively. Data of 15 different states are addressed in 1-15 columns and last column carry corresponding data of entire country.

|  | TN | KA | KL | UP | AP | MH | DL | GJ |
| --- | --- | --- | --- | --- | --- | --- | --- | --- |
| *Imported (from abroad)* | 13 | 33 | 96 | 0 | 3 | 8 | 3 | 3 |
| *State to state traveler* | 518 | 40 | 14 | 108 | 62 | 6 | 34 | 19 |
|  | MP | HR | JK | PB | RJ | TG | WB | India |
| Imported (from abroad) | 3 | 0 | 6 | 1 | 33 | 2 | 1 | 240 |
| State to state traveler | 0 | 17 | 6 | 12 | 26 | 18 | 0 | 938 |

**Figure 2:**
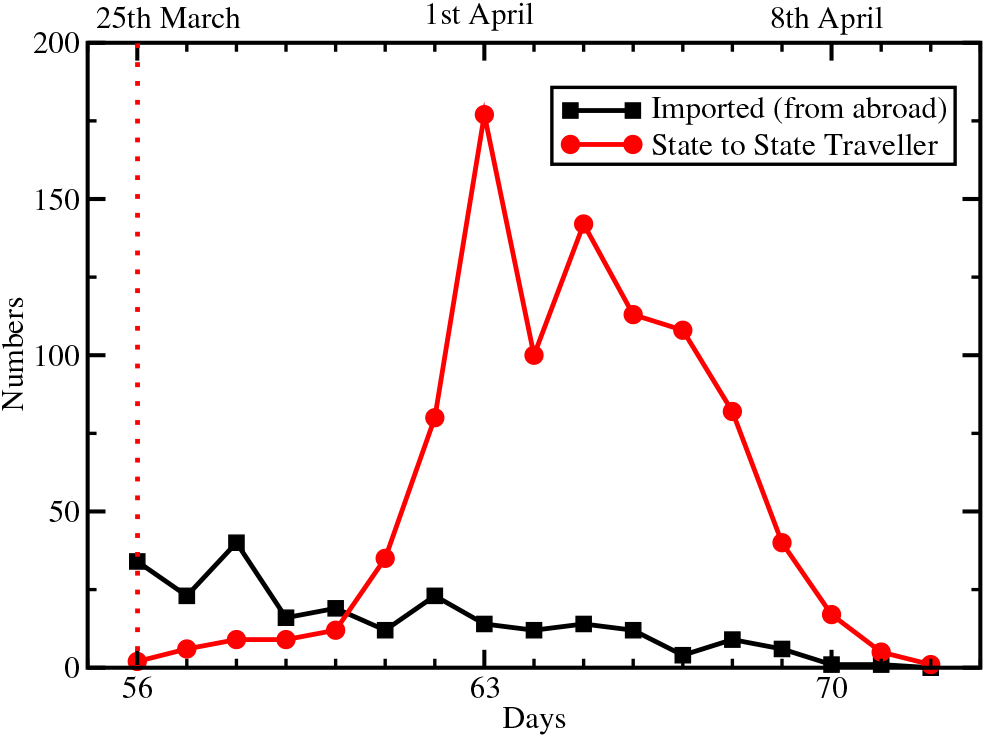
With increasing of days, the variation of new covid +ve cases, who came from abroad (black squares) and from outside state (red circles), marked as *Imported* (*from abroad*) and *State to state traveler* respectively.

### 3 5 states, having noticeable impact of HTs

The special characteristics of HTs data is that they are the traveler, who are appeared as +ve cases later and we can filtered out those during 2-3 weeks of lock down period. However, there should be some traveler, whose symptoms reveal instantly (within ∼ 24 − 48 hours) after reaching his/her own states. Separate these two type of imported cases (instantly and later detected cases) is a very challenging job as covid19 data collectors have to know the exact journey date of each imported cases. Interestingly, the separation become easy after 25^th^ March because of implementing lock-down. Analyzing the data roughly from 18 March (2 days before shutting down of airlines and rail transportation) to onwards, we will get instantly detected cases of imported group in the time-zone - *t* < 25 March. While later detected cases or HTs are located in the time-zone - *t* > 25 March. Now if we critically analyze and manipulate the data of HTs, then they might be shifted from lock down phase (≥ 25th March) to before lock-down (< 25th March) phase. If we exclude them then we might get little flatten curves of total cases. In this context, we have considered 5 states - Tamil Nadu (TN), Kerala (KL), Andhra Pradesh (AP), Uttar Pradesh (UP), Karnatak (KA), which carry noticeable amount of HTs and we noticed a change in the time profile of total cases and new cases when we go from including to excluding of HTs in the data. In Fig. (3), total cases (upper) and new cases (lower) data with (black circles) and without (pink triangles) HTs for TN (left), KL (right) are plotted. To mark the lock down period, we have drawn two vertical red dotted lines at 25th March and 3rd May. Now, we will attempt to fit the total cases data from 25th March to onwards via different possible Sigmoid functions with standard form [7, 8, 9, 3, 4]

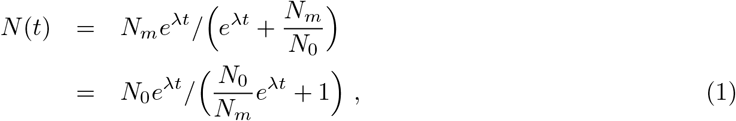

where *N*_0_ is initial value by setting 25th March as *t* = 0 and slope parameter λ and maximum values of total cases *N*_*m*_ will have to guess during fitting. For TN data, we sketch two Sigmoid curves with (1) *N*_*m*_ = 2, 000, λ = 0.3 and (2) *N*_*m*_ = 1, 700, λ = 0.25, represented by blue dash line and green dash-dotted line in left-upper and left-lower panel of Fig. (3). These two Sigmoid curves might be consider as lower and upper boundaries within which actual data of total cases take turning from rapid growth to flatten growth. This turning will be more visible in the new case vs days graphs. The new case can be grossly realized as time derivative of total cases. The time derivative of Sigmoid function follow the form [8, 9, 3, 4]

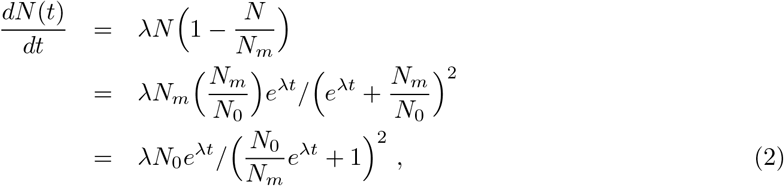

which first increases and later decreases with time. A peak in the new case data is expected around the days, where Sigmoid type total case data face the transformation from rapid growth to flatten growth. Last 19 days (31st March to 18th April) of TN new case data is almost saturated with a mild reduction. This time range may be considered as peak position of Sigmoid-type new case profile, expressed in Eq. (2). Corresponding time derivative of blue dash and green dash-dotted curves roughly show the pattern.

**Figure 3:**
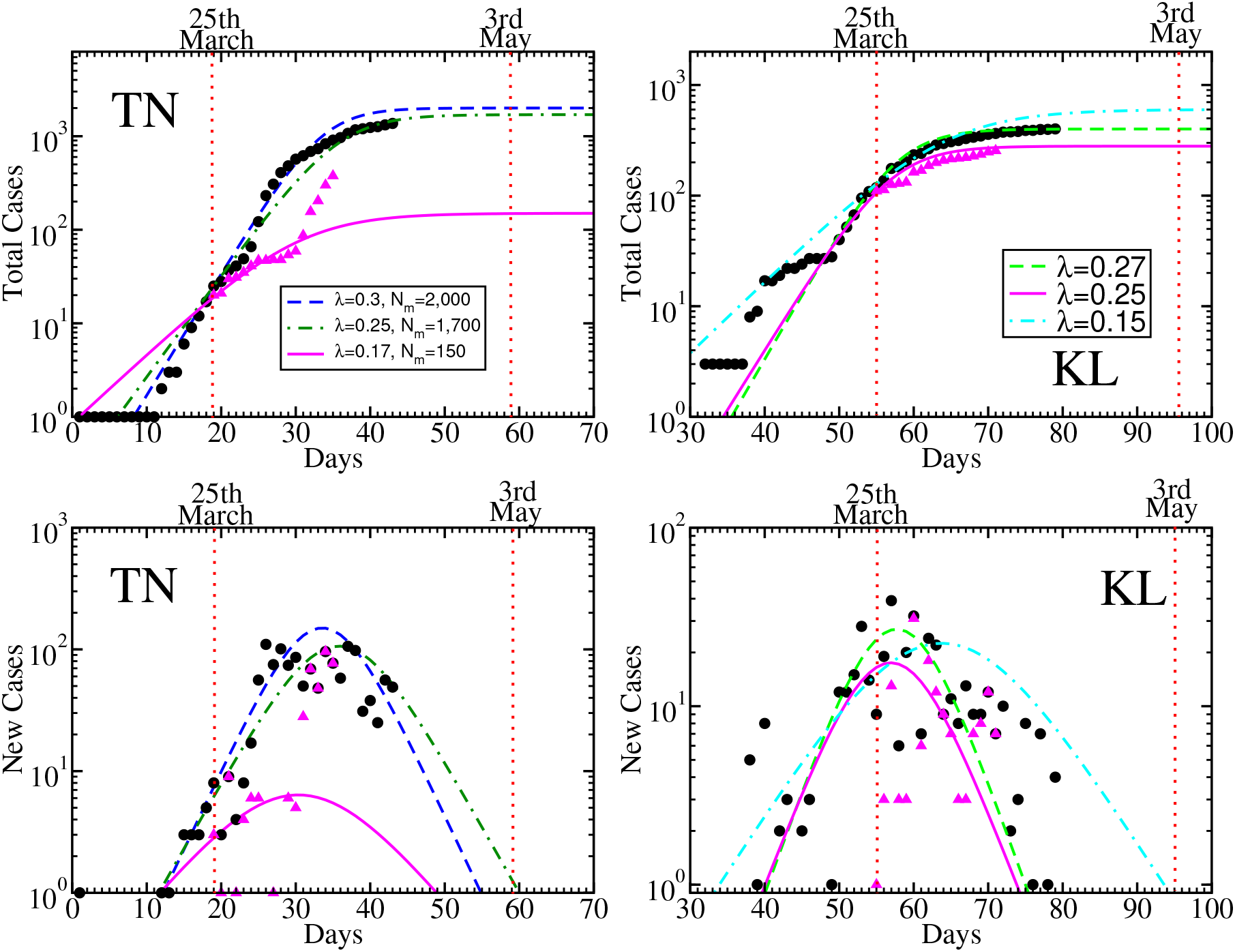
Total cases (upper), new cases (lower) data with (black circles) and without (pink triangles) hidden travelers (HTs), and their possible Sigmoid curves (different lines) of TN (left), KL (right). Two vertical red dotted lines are drawn at 25th March and 3rd May to mark the lock down period.

**Figure 4:**
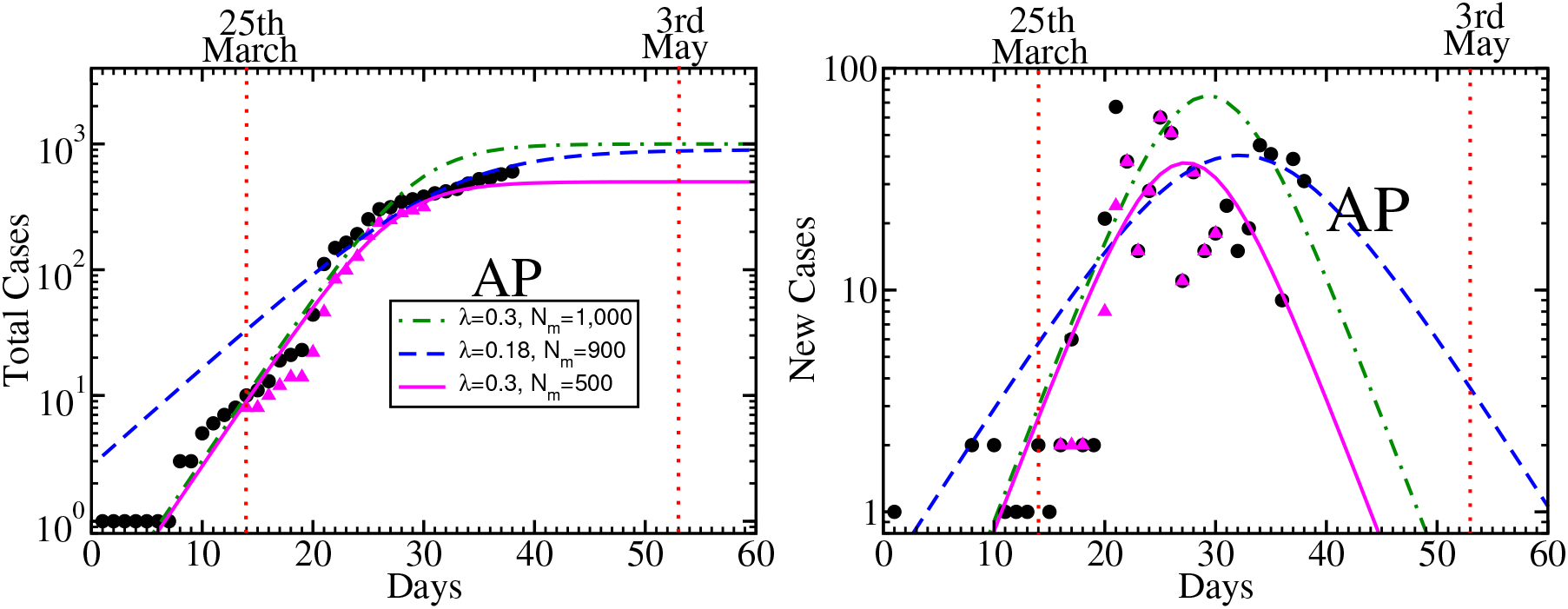
Same as earlier figures for AP.

**Figure 5:**
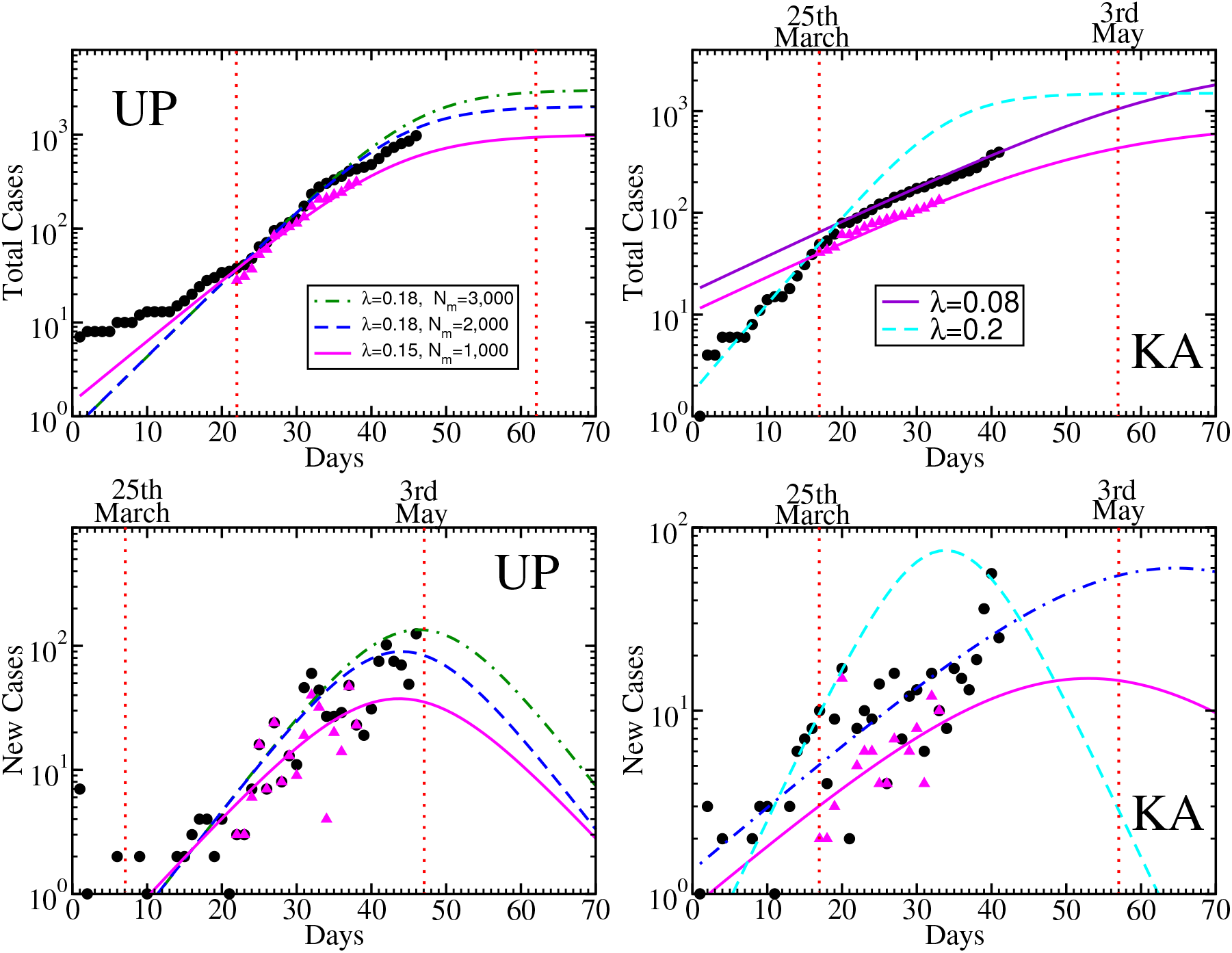
Same as earlier figures for UP (left) and KA (right).

**Figure 6:**
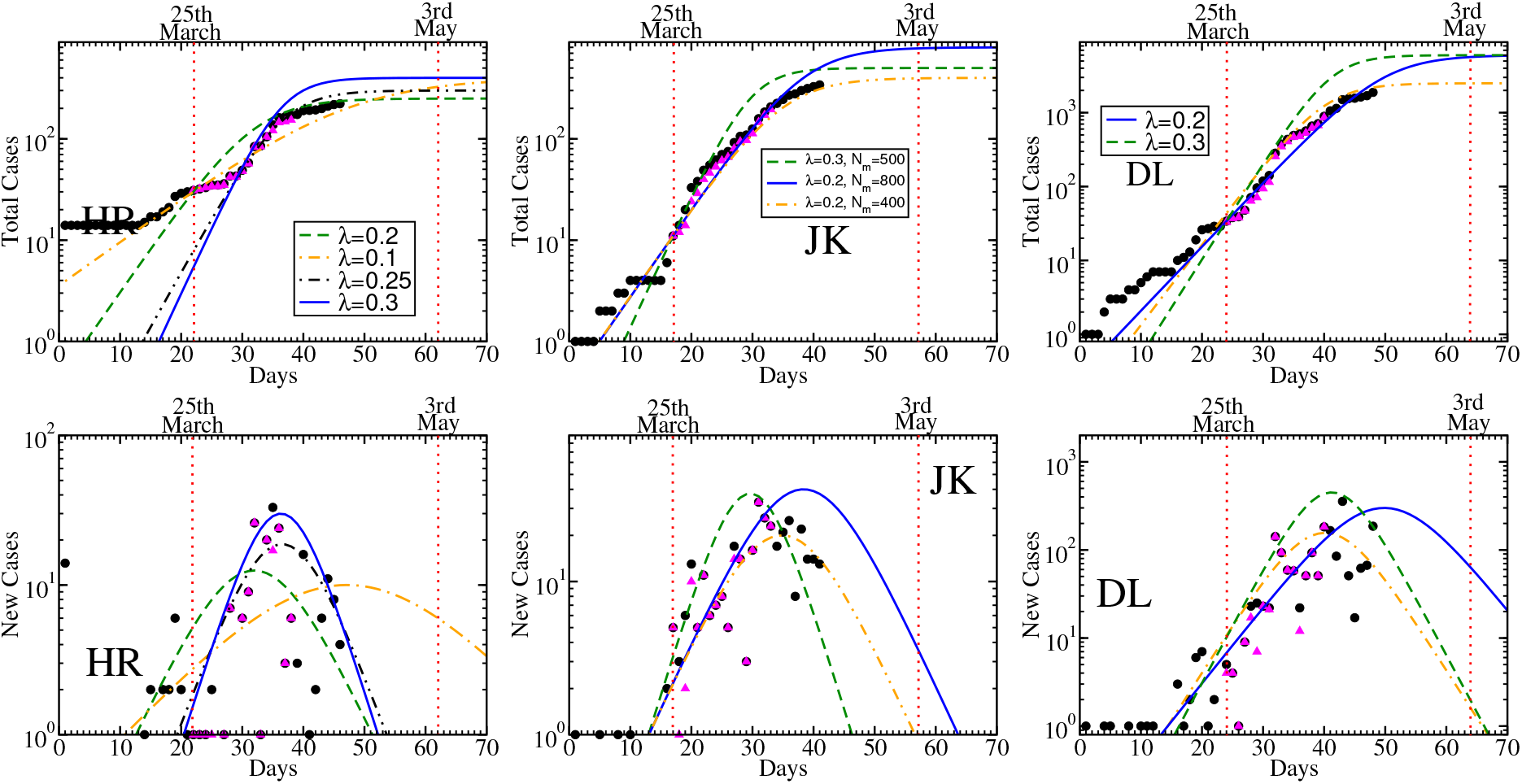
Same as earlier figures for HR (left), JK (middle) and DL (right).

**Figure 7:**
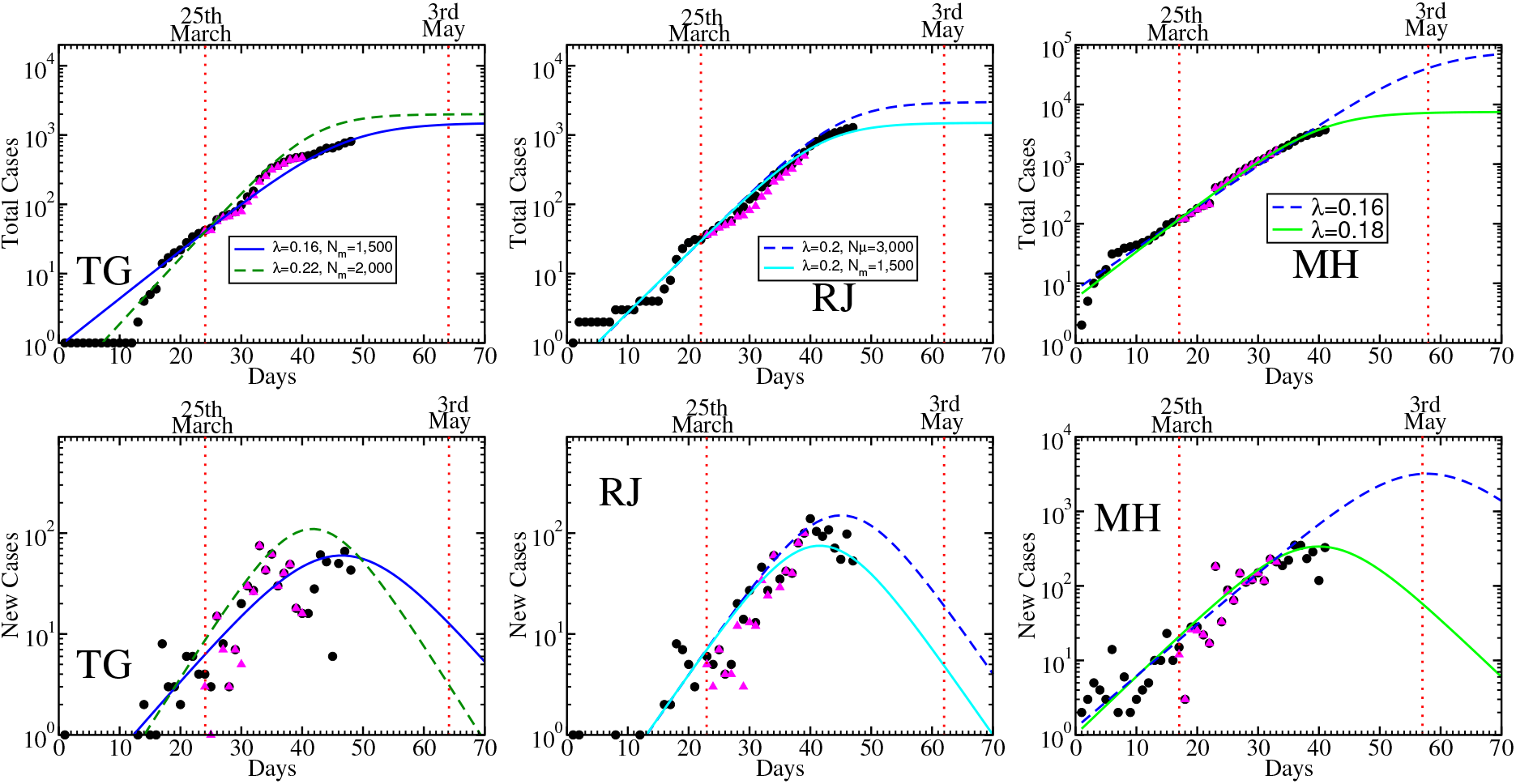
Same as earlier figures for TG (left), RJ (middle) and MH (right).

**Figure 8:**
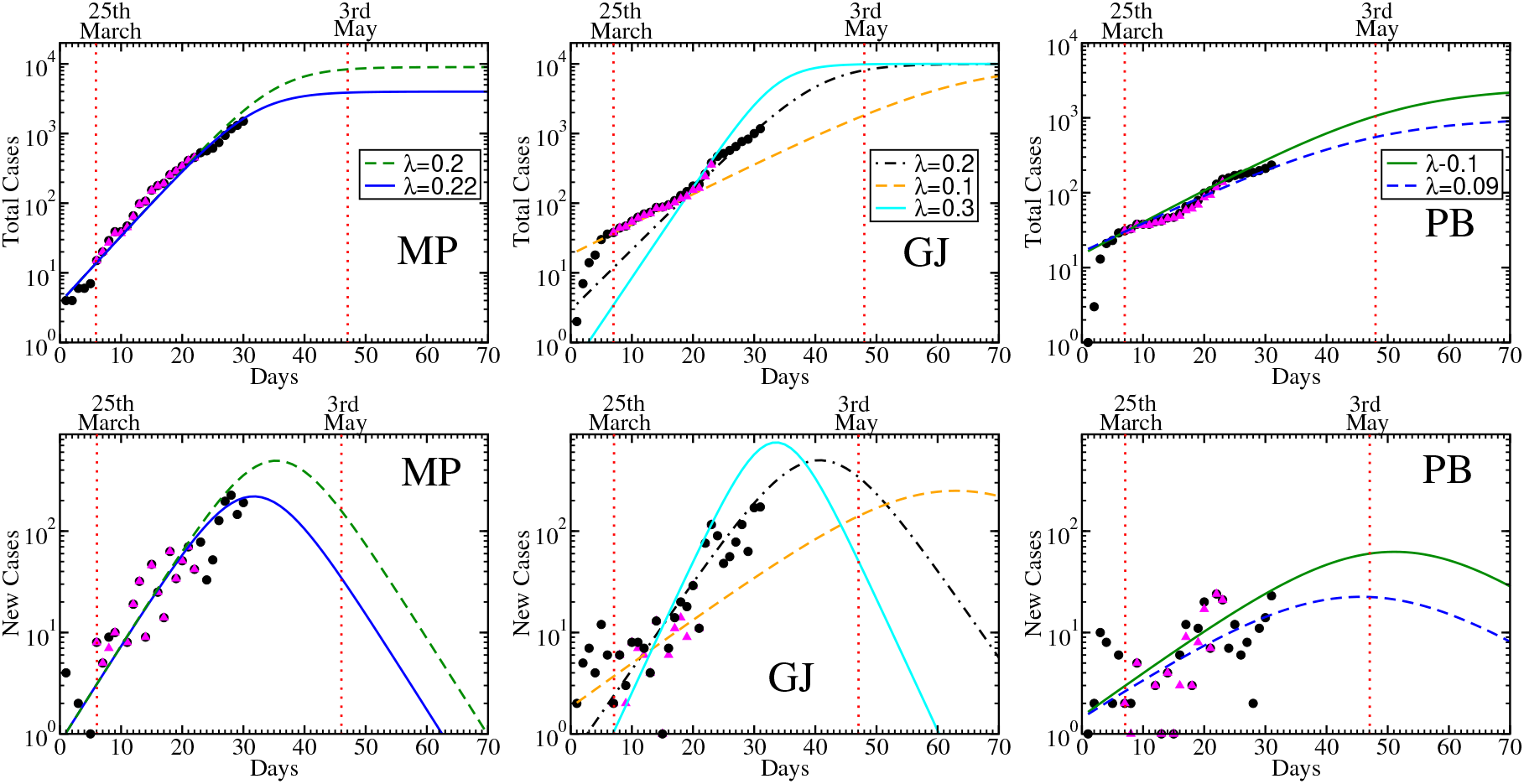
Same as earlier figures for MP (left), GJ (middle) and PB (right).

**Figure 9:**
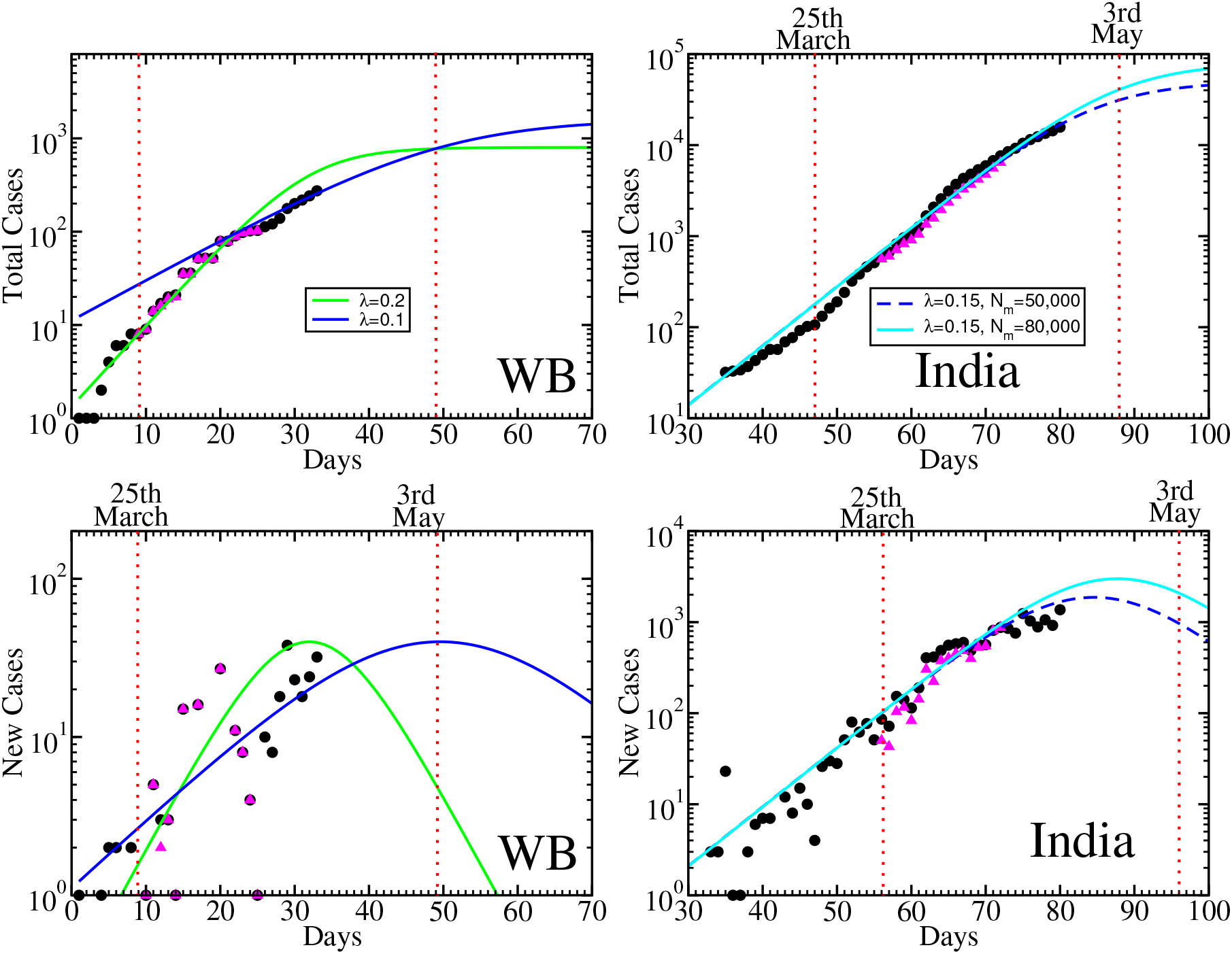
Same as earlier figures for WB (left), India (right).

Now, when we exclude HTs from total case data, then we find that pink triangles in left upper panel of Fig. (3) remain constant within the time zone 25th March to 5th April. It means that a large no of HTs during this period are responsible for getting enhanced actual data with larger growth slope. Now the excluded HT data (pink triangles) indicate a saturation trend in time zone 25th March to 5th April, which can be realized by a Sigmoid curve (pink solid line) with *N*_*m*_ = 170, λ = 0.17. Hence HTs of TN is responsible for the transformation of Sigmoid parameters *N*_*m*_ = 170 → *N*_*m*_ = 1, 700 − 2, 000 and λ = 0.17 → λ = 0.25 − 0.3.

In brief, we can summarize the Sigmoid-type trend of present data during the lock-down period in bullet points.

#### 3.1 TN

- Including HTs, (Optimistic) saturation value of total cases might be *N*_*m*_ = 1, 700-2, 000.
- Including HTs, if we assume (optimistically) the peak value of new cases around 8^th^-10^th^ April, then new cases might be reduced to marginal on 28^th^ April-4^th^ May.
- Excluding HTs, (Optimistic) saturation value of total cases might be *N*_*m*_ = 150.
- Excluding HTs, if we assume (optimistically) the peak value of new cases around 5^th^ April, then new cases might be reduced to marginal on 23^rd^ April.

For remaining states and union territories, we have followed similar kind of sketching, hence instead of elaborate discussions of their corresponding graphs, main bullet points will be addressed.

#### 3.2 KL

- Including HTs, (Optimistic) saturation value of total cases might be *N*_*m*_ = 400-600.
- Including HTs, if we assume (optimistically) the peak value of new cases around 27^th^ March-2^nd^ April, then new cases might be reduced to marginal on 15^th^ April-2^nd^ May.
- Excluding HTs, (Optimistic) saturation value of total cases might be *N*_*m*_ = 300.
- Excluding HTs, if we assume (optimistically) the peak value of new cases around 27^th^ March, then new cases might be reduced to marginal on 13^th^ April.

#### 3.3 AP

- Including HTs, (Optimistic) saturation value of total cases might be *N*_*m*_ = 900-1, 000.
- Including HTs, if we assume (optimistically) the peak value of new cases around 9^th^-12^nd^ April, then new cases might be reduced to marginal on 28^th^ April-12^nd^ May.
- Excluding HTs, (Optimistic) saturation value of total cases might be *N*_*m*_ = 500.
- Excluding HTs, if we assume (optimistically) the peak value of new cases around 7^th^ April, then new cases might be reduced to marginal on 23^th^ April.

#### 3.4 UP

- Including HTs, (Optimistic) saturation value of total cases might be *N*_*m*_ = 2, 000-3, 000.
- Including HTs, if we assume (optimistically) the peak value of new cases around 30^th^ April-11^th^ May, then new cases might be reduced to marginal on 1^st^-5^th^ June.
- Excluding HTs, (Optimistic) saturation value of total cases might be *N*_*m*_ = 2, 000.
- Excluding HTs, if we assume (optimistically) the peak value of new cases around 30^th^ April, then new cases might be reduced to marginal on 1^st^ June.

#### 3.5 KA

- Including HTs, (Optimistic) saturation value of total cases might be *N*_*m*_ = 1, 500-2, 000.
- Including HTs, if we assume (optimistically) the peak value of new cases around 10^th^ April-11^th^ May, then new cases might be reduced to marginal on 8^th^ April-18^th^ July.
- Excluding HTs, (Optimistic) saturation value of total cases might be *N*_*m*_ = 700.
- Excluding HTs, if we assume (optimistically) the peak value of new cases around 30^th^ April, then new cases might be reduced to marginal on 19^th^ June.

At the end of this section, if we collect the outcomes, then we can visualize the impact of HTs in new and total case data, which clearly show that lower *N*_*m*_ in total cases and peak, marginal positions in new cases are shifted to larger values because of HTs.

### 4 10 states, having ignorable impact of HTs

Apart from the 5 states, described above, we have also repeated our analysis to 10 more states, Haryana (HR), Jammu & Kashmir (JK), Delhi (DL), Telangana (TG), Maharashtra (MH), Madhya Pradesh (MP), Gujrat (GJ), Punjab (PB), West Bengal (WB), in where inclusion or exclusion of HTs does not make any noticeable change. So here, we only two bullet points for each states instead of four. These are documented below.

#### 4.1 HR

- *N*_*m*_ = 250-400.
- Peak positions in new cases : 5^th^-20^th^ April; Position(s) of marginal new cases : 22^th^ April-23^th^ May.

#### 4.2 JK

- *N*_*m*_ = 400-800.
- Peak positions in new cases : 6^th^-16^th^ April; Position(s) of marginal new cases : 22^nd^ April-9^th^ May.

#### 4.3 DL

- *N*_*m*_ = 2, 500-6, 000.
- Peak positions in new cases : 10^th^-20^th^ April; Position(s) of marginal new cases : 6^th^-25^th^ May.

#### 4.4 TG

- *N*_*m*_ = 1, 500-2, 000.
- Peak positions in new cases : 11^th^-17^th^ April; Position(s) of marginal new cases : 9^th^-19^th^ May.

#### 4.5 RJ

- *N*_*m*_ = 1, 500-3, 000.
- Peak positions in new cases : 12^th^-16^th^ April; Position(s) of marginal new cases : 11^th^-18^th^ May.

#### 4.6 MH

- *N*_*m*_ = 7, 500-70, 000.
- Peak positions in new cases : 18^th^ April-4^th^ May; Position(s) of marginal new cases : 25^th^ May-1^st^ July.

#### 4.7 MP

- *N*_*m*_ = 4, 000-9, 000.
- Peak positions in new cases : 19^th^-23^th^ April; Position(s) of marginal new cases : 18^th^-27^th^ May.

#### 4.8 GJ

- *N*_*m*_ = 7, 000-10, 000.
- Peak positions in new cases : 20^th^ April-25^th^ May; Position(s) of marginal new cases : 16^th^ May-27^th^ July.

#### 4.9 PB

- *N*_*m*_ = 1, 000-2, 500.
- Peak positions in new cases : 3^th^ May-8^th^ May; Position(s) of marginal new cases : 20^th^ June-2^th^ July.

#### 4.10 WB

- *N*_*m*_ = 800-1, 400.
- Peak positions in new cases : 18^th^ April-6^th^ May; Position(s) of marginal new cases : 11^th^ May- 24^th^ June.

#### 4.11 India

- *N*_*m*_ = 50, 000-80, 000.
- Peak positions in new cases : 23^rd^ April-25^th^ April; Position(s) of marginal new cases : 22^nd^ June-27^th^ June.

At the end, if we summarize outcomes of earlier two section, then our selective 15 states can be classified roughly into three categories -

1. The states, whose data face peak or saturation in new cases and then a visible reduction, can be thought to be in good situation. KL and HR belong to this category.
2. The states, whose increasing data face a saturation (with a certain error bar) during 5-10 days in new cases and last data point become smaller than saturation/peak value, can be said to be in intermediate situation. JK, TN, AP, TG, RJ, MH belong to this category.
3. The states, whose increasing data don’t reveal any saturation trend yet, can be considered to be in bad situation. UP, KA, PB, GJ, MP, WB belong to this category.

Among the example of 2nd category, JK, TN, AP can be considered toward first category as their last data points exhibit some reduction. Their extrapolation hopes that they might be in first category before ending of ongoing lock down (3rd May). On the other hand TG, RJ, MH presently pass through a saturation value. Until we don’t see any visible reduction in future data, it is little hard to predict. So they are quite close to third category. For third category states - UP, KA, PB, GJ, MP, WB, predicting something is really a hard task. In this regards, our prediction can be considered as a boundary of best optimistic possibilities, but its opposite boundary might be quite disaster.

## 5 Summary and Interpretation

The present article is mainly focused on imported covid19 data of India and extracted few interesting points. The summary of the investigating steps are as follows. First, we sketch the pattern of imported data with days in new and total cases. Defining the ratio between imported to all covid19 cases, we have found its variation with time. Since Covid19 is an imported disease for all country (except China), so the ratio should be one in the beginning and later it will be reduced. For covid19 data of India, we find that from 30th January to 4th March the ratio remain one and after that, the ratio started to reduce, which implies that imported person have started to spread infection to others. During 4th March to 20/25th March the reduced value of ratio remain within 0.6 to 0.8 but after that period it face gradual reductions due to effect of lock down from 25th March to onwards.

The imported cases, documented after 25th March, probably traveled before 20th March, which means that their covid19 symptom take time to reveal. For our convenience, we have called them as HTs (hidden travelers). We explored the impact of HTs on Sigmoid type pattern of total and new cases data. In general, the Sigmoid profile of total cases transform from rapid growth to flatten growth and saturates towards a maximum value *N*_*m*_. On the other hand, it’s new case profile increase, then saturate to a peak value(s) and then decrease to marginal values. We have found that this maximum value *N*_*m*_ in total case, and peak, marginal positions in new cases are increased because of the HTs. This enhancing factor will not work further after 8th April, from when the data of HTs is completely disappeared. This might be considered as a quantitative factor, reflecting a straight forward benefit of lock down. However it has been noticed in 5 states - TN, KL, AP, KA, UP.

After that, we have also repeated our Sigmoid analysis for 10 more states - HR, JK,DL,TG, MH, MP, GJ, PB and WB. Analyzing the Sigmoid profile of all 15 states, they may be classified as three categories, depending upon latest data of new case with respect to the peak value of Sigmoid functions. First category states like KL, HR crosses the peak position and they turned towards the marginal values. second category states like JK, TN, AP, TG, RJ, MH probably just reach the peak position, which can be realized from the last saturation trends in their new case data. Last category states like UP, KA, PB, GJ, MP and WB are in before the peak position. Based on our (optimistic) predicted curves, KL, HR, JK, TN and AP might be reached to marginal values within 3rd May (end of second phase lock down) but the reamining of 15 states might need a third phase of lock down or alternative preventive measures.

Before going to any relaxation plan of lock down, severity of state to state transfer data should be keep in mind. From 30th January to 8th April, total imported cases are 1,533 among which 595 were imported from abroad and 938 were from state to state transferred case. Interestingly this 938 numbers is identified within 25th March to 8th April. So one can guess that this state to state transfer crowed occurred within 18th to 20th March who were diagnosed later from 25th March to 8th April. From here we can roughly estimate 1,000 positive cases, transfered from other state if transportation will be reopened for two days. They will further continue their second generation spreading before detected as +ve case. So, from present study, we may visualize a future second wave growth, coming from re-birthing imported data through lock-down/transportation relaxation. Maintaining state to state transportation blocking might be appeared as very promising solution. For more critical solution, one can go through the same way of investigations for different zones of each states.

## Data Availability

Cited in references.....

https://www.covid19india.org/

## Acknowledgment

SM and SG thank to their daughter Adrika Ghosh for allowing time for this investigation during lock-down period.

